# Neural substrates in Parkinson’s Disease psychosis: A systematic review

**DOI:** 10.1101/2022.12.23.22283889

**Authors:** Sara Pisani, Brandon Gunasekera, Yining Lu, Miriam Vignando, Dominic ffytche, Dag Aarsland, K. Ray Chaudhuri, Clive Ballard, Jee-Young Lee, Yu Kyeong Kim, Latha Velayudhan, Sagnik Bhattacharyya

## Abstract

Neural underpinnings of Parkinson’s Disease psychosis (PDP) remain unclear to this day with relatively few studies and reviews available. Using a systematic review approach, here we aimed to qualitatively synthesize evidence from studies investigating PD psychosis-specific alterations in brain structure, function or chemistry using different neuroimaging modalities. PubMed, Web of Science and Embase databases were searched for fMRI, rsfMRI, DTI, PET, and SPECT studies comparing PDP patients with PD patients without psychosis (PDnP). We report findings from 18 studies (291 PDP patients, mean age ± SD = 68.65 ± 3.76 years; 48.5% males; 433 PDnP patients, mean age ± SD = 66.97 ± 3.80 years; 52% males). Qualitative synthesis revealed widespread patterns of altered brain function across task-based and resting-state fMRI studies in PDP compared to PDnP patients. Similarly, white matter abnormalities were reported in parietal, temporal, and occipital regions. Hypometabolism and reduced dopamine transporter binding were also reported whole brain and in subcortical areas. This suggests extensive alterations affecting regions involved in high order visual processing and attentional networks.

## 1. Introduction

Parkinson’s Disease (PD) is one of the most common neurodegenerative conditions worldwide, with an estimate of 9.3 million people suffering from it by 2030 (Dorsey et al., 2007). Although PD is mostly recognised as a movement disorder characterised by bradykinesia, tremors, rigidity and postural instability, there are in addition several non-motor symptoms (e.g., depression, constipation and urine problems, sleep issues, and many others) that negatively impact on the patient’s quality of life (Aarsland et al., 2013; Antonini et al., 2018; Balestrino & Martinez-Martin, 2017; Chaudhuri et al., 2006). Specifically, PD patients may experience hallucinations and delusions, which are collectively referred to as PD psychosis (PDP), which are distressing, debilitating and associated with increase in care burden and risk of hospitalisation (Aarsland et al., 2007; Ffytche et al., 2017; Martinez-Martin et al., 2015). Severity and duration of PD, dopaminergic medications, sleep disorders, cognitive decline, widespread Lewy Body pathology, and late PD onset can be risk factors for developing psychosis symptoms (Barrett et al., 2018; Chang & Fox, 2016; Factor et al., 2014; Ffytche et al., 2017; Williams & Lees, 2005).

Evidence from individual studies employing different neuroimaging modalities have started to uncover the mechanisms underlying the emergence of psychotic symptoms in PD. Results from structural magnetic resonance imaging (MRI) studies have reported whole-brain cortical volume loss (Bejr-Kasem et al., 2019; Gama et al., 2014; Ramírez-Ruiz et al., 2007) in PDP compared to PD patients without psychosis (PDnP), and especially in regions encompassing the ventral and dorsal visual pathways, the parietal, temporal and occipital lobe (Pisani et al., 2022), as well as hippocampus (Pezzoli et al., 2019; Pezzoli et al., 2021; Vignando et al., 2022). Complementing this, task-based and resting state functional MRI (fMRI) studies have reported altered temporo-parietal-occipital activation and connectivity and research using positron emission tomography (PET) and diffusion tensor imaging (DTI) has also observed abnormal metabolism and structural connectivity in large-scale brain networks respectively (Boecker et al., 2007; Hall et al., 2019; Hepp et al., 2017; Lefebvre et al., 2016; Shine, Halliday, et al., 2014; Shine et al., 2015; Stebbins et al., 2004). Most reviews on the topic have focused on structural correlates involved in PDP (Alzahrani & Venneri, 2015), and few have examined its neurofunctional substrates (Lenka et al., 2015). Integrating evidence from different neuroimaging modalities may help unravel altered mechanisms in PD psychosis, though this may be a challenging undertaking due to the different techniques and methodologies. As a number of recent reviews on the structural correlates of PDP exist (Pezzoli et al., 2021; Vignando et al., 2022) including one from our group (Pisani et al., 2022), here we primarily focused on summarizing evidence from studies using other neuroimaging modalities. In particular, we have endeavoured to qualitatively synthesise evidence from separate studies employing a range of neuroimaging modalities (other than structural MRI) such as task-based and resting-state fMRI, PET and DTI. The aim of this review was to summarise the main patterns of abnormal activation, connectivity and metabolism in PD psychosis compared to PD patients without psychosis, to provide a more comprehensive account of existing evidence from single separate studies regarding the neural correlates of PD psychosis that may complement evidence synthesized from structural MRI studies.

## 2. Methods

### 2.1. Search strategy and eligibility criteria

We followed the Preferred Reporting Items for Systematic Reviews and Meta-Analyses (PRISMA) guidelines (Moher et al., 2009) (PROSPERO registration number: CRD42020221904) and a detailed description of the search strategy is outlined in Supplementary Material 1. We systematically searched PubMed, Web of Science, Embase and the Neurosynth database and we included studies if they examined brain alterations associated with psychosis symptoms (i.e., hallucinations and/or delusions) after PD diagnosis and if they used neuroimaging techniques (i.e., functional MRI (fMRI), resting-state fMRI (rsfMRI), positron emission tomography (PET), single photon emission computerised tomography (SPECT), diffusion tensor imaging (DTI), magnetic resonance spectroscopy (MRS)) and a case-control design.

### 2.2. Data extraction and synthesis

Data extraction involved: study details (e.g., authors’ names, year of publications), study design, neuroimaging modality as well as scanner characteristics (e.g. manufacturer), sample size, sample characteristics (e.g. age, gender, education levels), PD onset age, disease duration, PD symptoms, clinical measures of psychosis symptoms, cognitive measures, PD medications (expressed in Levodopa equivalent daily dose (LEDD)), list of any other concurrent medications, and any other clinical outcome measures (e.g., depression). Data extraction was conducted independently by two researchers (SP, YL) with discrepancies addressed through consensus or discussion with senior researchers. The purpose of this review was to specifically examine evidence from neuroimaging modalities other than structural MRI approach. Therefore, we focused on task-based fMRI, resting-state fMRI (rsfMRI), PET, SPECT and DTI studies. Due to the lack of comparable methodological approaches (Müller et al., 2018) (e.g., tract-based spatial statistics vs. fractional anisotropy for DTI studies; whole-brain analysis vs. ROI analysis) that would allow meaningful synthesis of data we synthesised these studies in a qualitative manner. Results were specifically reviewed to compare PDP and PDnP patients. Studies that included multiple neuroimaging modalities were considered as individual studies for each modality they used. To facilitate the qualitative synthesis of evidence from the included studies, we represented visually the cortical and sub-cortical areas with patterns of abnormalities in PDP patients using *ggseg* (Mowinckel & Vidal-Piñeiro, 2020) on R (version 4.0.3) (Team, 2013) and formatted using the Desikan-Killiany brain atlas (Desikan et al., 2006).

### 2.3. Quality rating assessment

The quality of the studies included was assessed with the Newcastle-Ottawa rating Scale for case-control studies (Wells et al., 2000) which assesses three methodological domains using a star system: “Selection” (i.e., definition and selection of cases and controls, maximum of one star), “Comparability” (i.e., comparison of cases and controls on the variables of interest and covariates, maximum of two stars), and “Exposure” (i.e., definition of the exposure or condition, maximum of one star). In this review, PDP patients were compared to PDnP patients, thus the latter acted as a control or comparison group.

## 3. Results

### 3.1. Study characteristics

Our systematic search identified 12 studies (Bejr-Kasem et al., 2019; Hepp et al., 2017; Kiferle et al., 2014; Lee et al., 2016; Lee et al., 2017; Nishio et al., 2018; Park et al., 2013; Shine et al., 2015; Stebbins et al., 2004; Yao et al., 2014; Yao et al., 2016; Zarkali et al., 2020), with an additional 5 studies (Boecker et al., 2007; Firbank et al., 2018; Jaakkola et al., 2017; Knolle et al., 2020; Lefebvre et al., 2016) identified from systematic reviews and one (Meppelink et al., 2009) from the Neurosynth database. Thus, we included a total of 18 published articles (Fig. 1) reporting on a total of n=724 patients, of which 291 had psychosis symptoms (mean age ± SD = 68.65 ± 3.76 years; 48.5% males). 433 PDnP patients did not have such symptoms (mean age ± SD = 66.97 ± 3.80 years; 52% males) and acted as the ‘control group’ (Table 1). Of the 18 studies, two studies included multimodal neuroimaging design (Firbank et al., 2018; Yao et al., 2016) and each modality was considered as an independent study. Full quality ratings can be found in Supplementary Material 2 (eTable1).

**Fig. 1.**
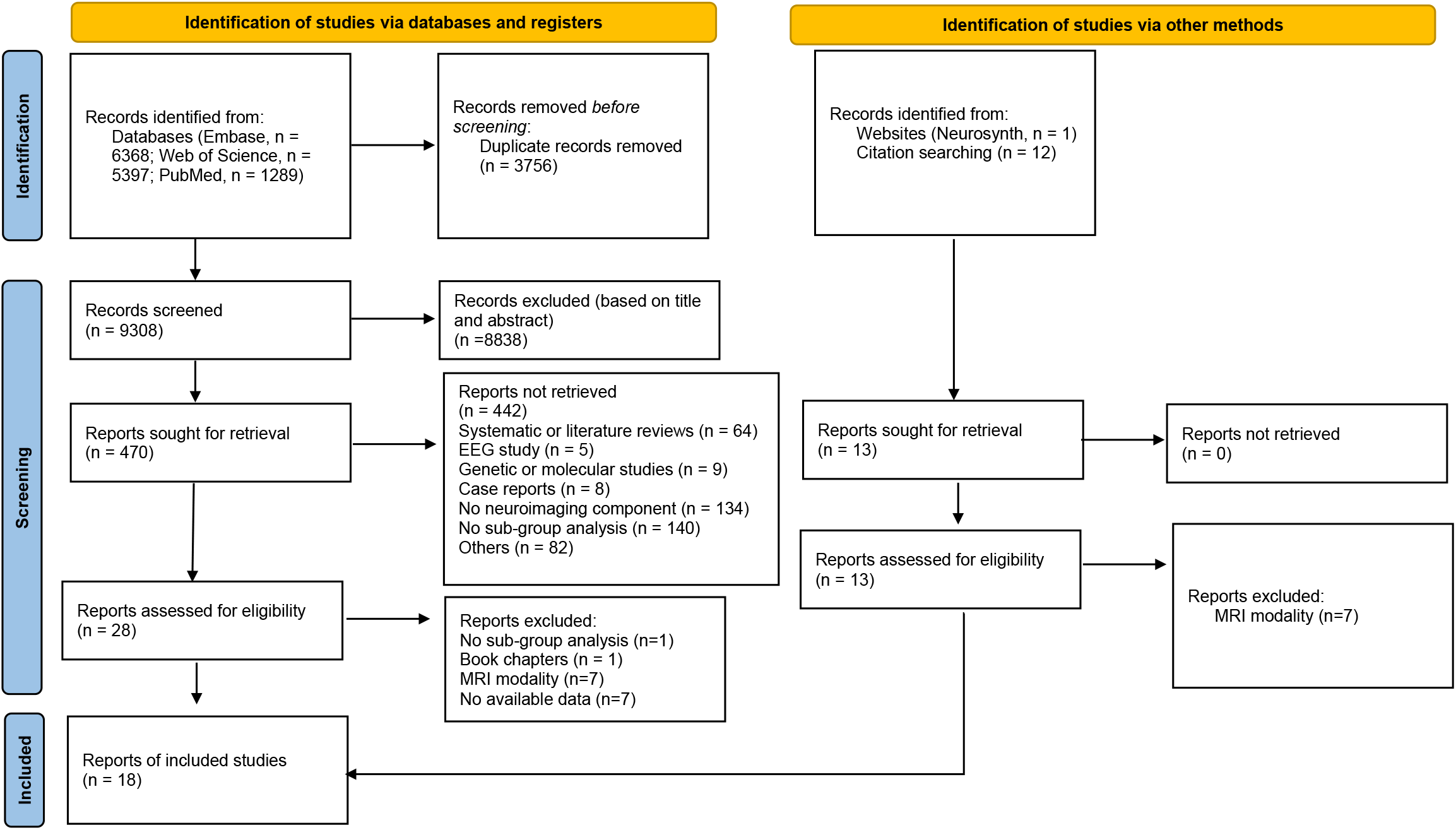
PRISMA flowchart with the study selection procedures.

**Table 1.**
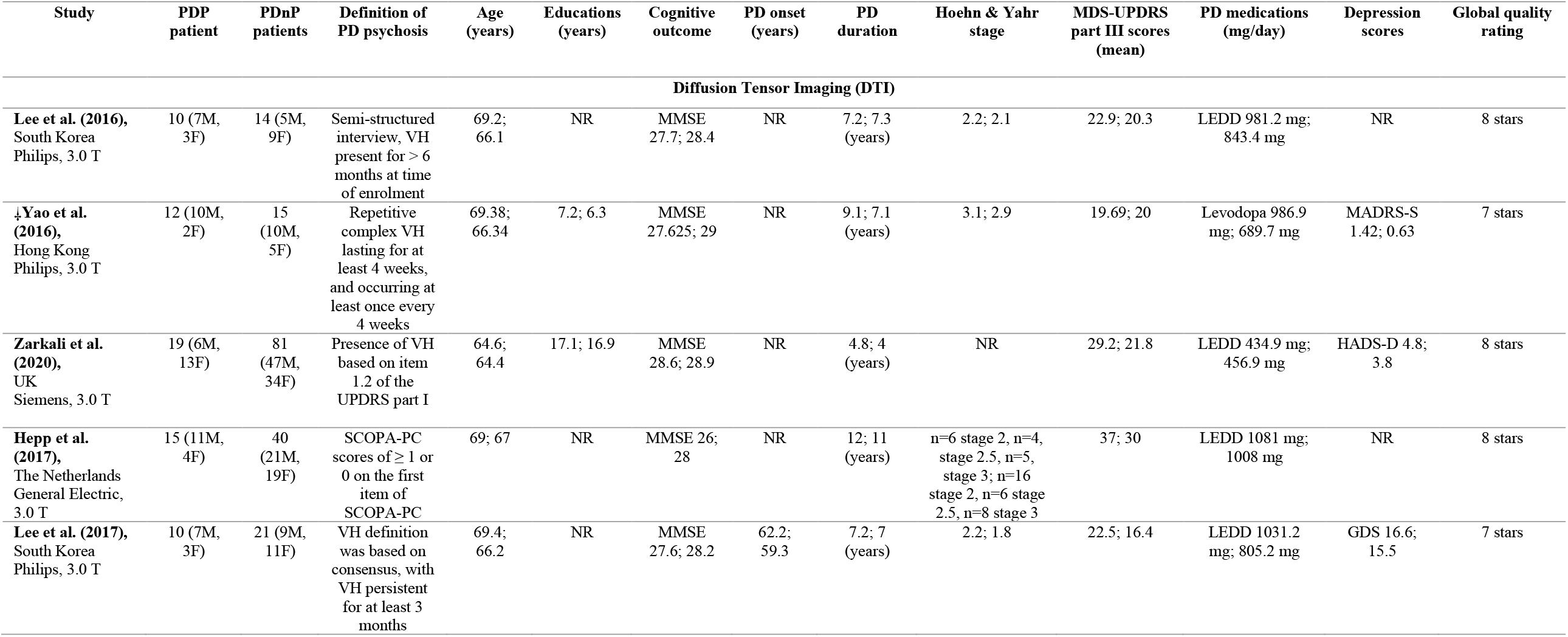
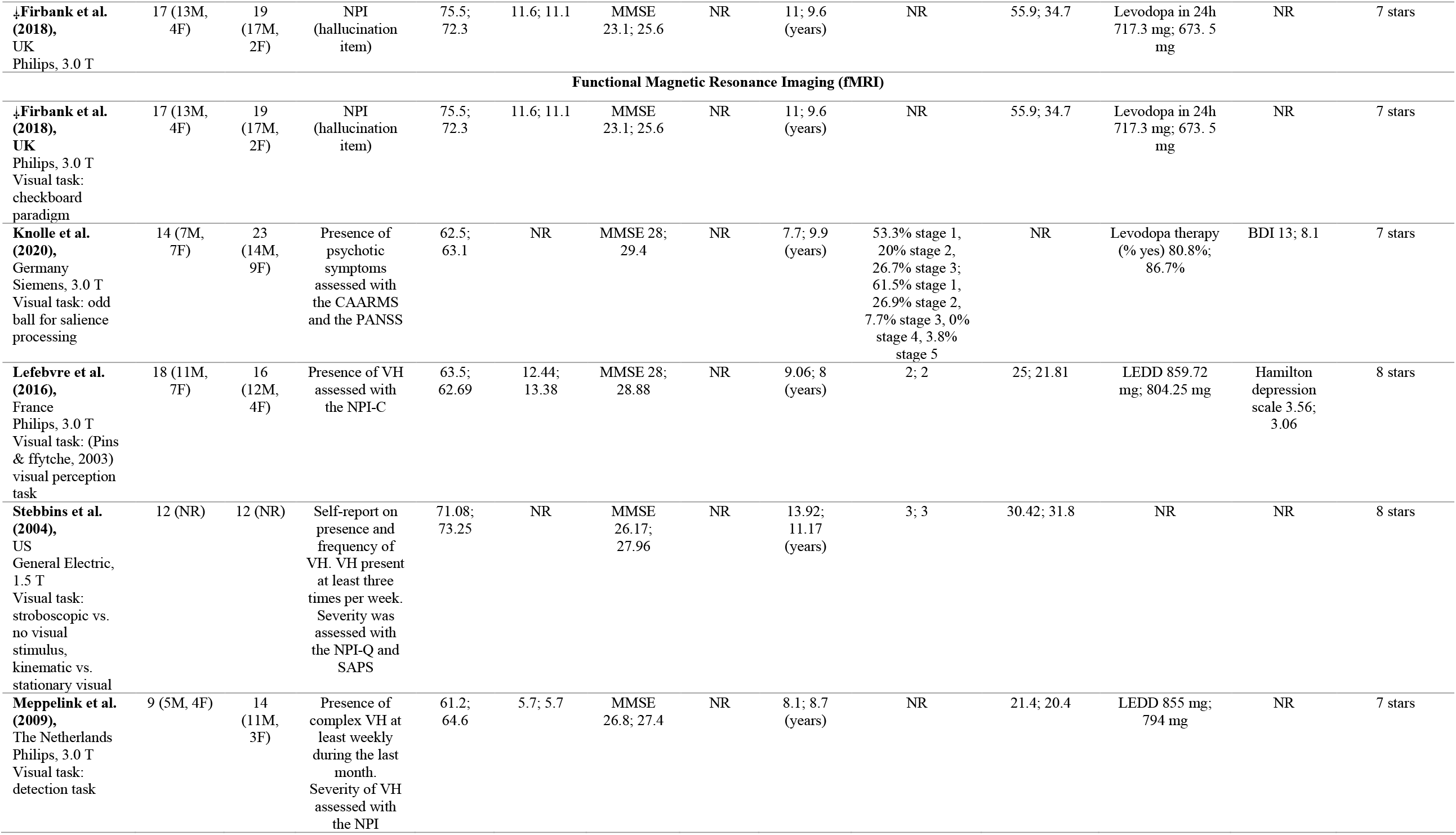
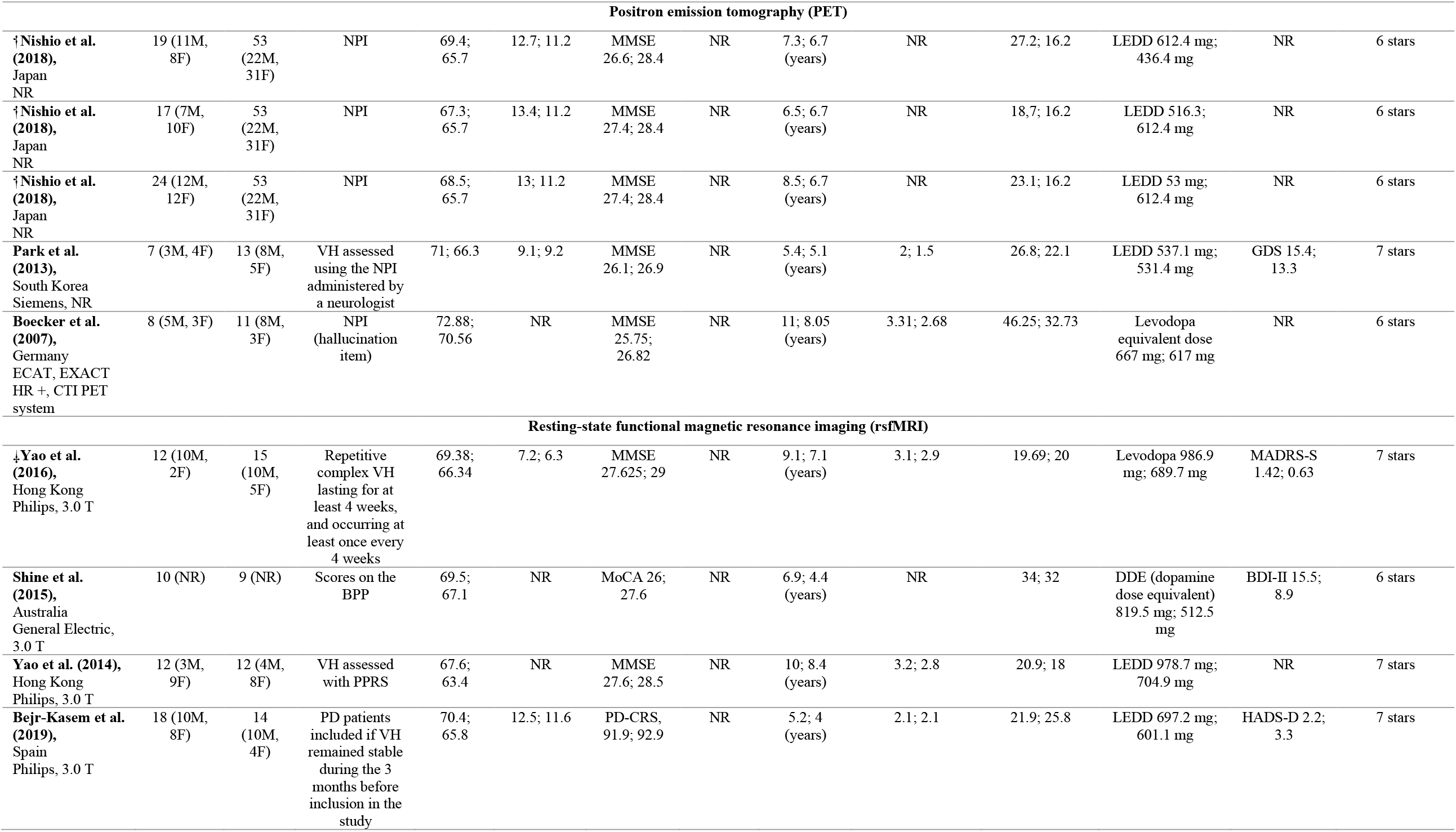
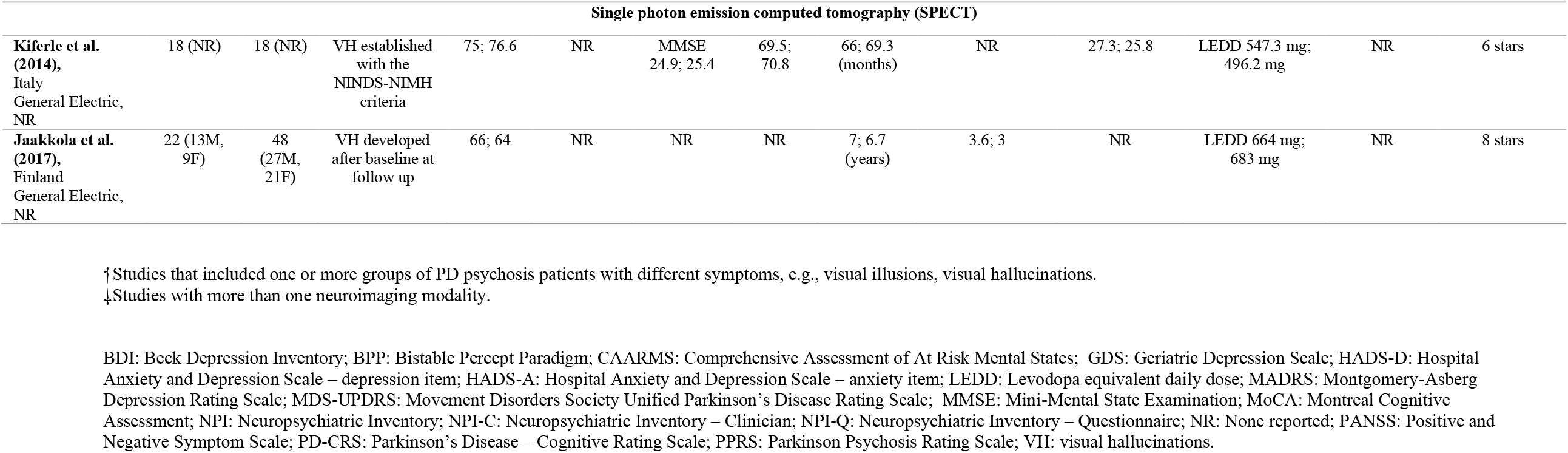
Study description (including location and scanner details) with statistics (mean, unless otherwise specified) presented for each patient group (PDP patients; PDnP patients). Number of patients for each group are presented alongside gender (males, M; females, F), years of education, scores on cognitive outcomes. Clinical variables of interest are also reported, e.g., motor symptoms and stage (according to the MDS-UPDRS and Hoehn and Yahr stages), PD medications (expressed in mean daily mg where available), and depression scores where available, and global quality assessments for each study.

| Study | PDP patient | PDnP patients | Definition of PD psychosis | Age (years) | Educations (years) | Cognitive outcome | PD onset (years) | PD duration | Hoehn & Yahr stage | MDS-UPDRS part III scores (mean) | PD medications (mg/day) | Depression scores | Global quality rating |
| --- | --- | --- | --- | --- | --- | --- | --- | --- | --- | --- | --- | --- | --- |
| **Diffusion Tensor Imaging (DTI)** |  |  |  |  |  |  |  |  |  |  |  |  |  |
| **Lee et al. (2016)**, South Korea Philips, 3.0 T | 10 (7M, 3F) | 14 (5M, 9F) | Semi-structured interview, VH present for > 6 months at time of enrolment | 69.2; 66.1 | NR | MMSE 27.7; 28.4 | NR | 7.2; 7.3 (years) | 2.2; 2.1 | 22.9; 20.3 | LEDD 981.2 mg; 843.4 mg | NR | 8 stars |
| **Yao et al. (2016)**, Hong Kong Philips, 3.0 T | 12 (10M, 2F) | 15 (10M, 5F) | Repetitive complex VH lasting for at least 4 weeks, and occurring at least once every 4 weeks | 69.38; 66.34 | 7.2; 6.3 | MMSE 27.625; 29 | NR | 9.1; 7.1 (years) | 3.1; 2.9 | 19.69; 20 | Levodopa 986.9 mg; 689.7 mg | MADRS-S 1.42; 0.63 | 7 stars |
| **Zarkali et al. (2020)**, UK Siemens, 3.0 T | 19 (6M, 13F) | 81 (47M, 34F) | Presence of VH based on item 1.2 of the UPDRS part I | 64.6; 64.4 | 17.1; 16.9 | MMSE 28.6; 28.9 | NR | 4.8; 4 (years) | NR | 29.2; 21.8 | LEDD 434.9 mg; 456.9 mg | HADS-D 4.8; 3.8 | 8 stars |
| **Hepp et al. (2017)**, The Netherlands General Electric, 3.0 T | 15 (11M, 4F) | 40 (21M, 19F) | SCOPA-PC scores of $\geq 1$ or 0 on the first item of SCOPA-PC | 69; 67 | NR | MMSE 26; 28 | NR | 12; 11 (years) | n=6 stage 2, n=4, stage 2.5, n=5, stage 3; n=16 stage 2, n=6 stage 2.5, n=8 stage 3 | 37; 30 | LEDD 1081 mg; 1008 mg | NR | 8 stars |
| **Lee et al. (2017)**, South Korea Philips, 3.0 T | 10 (7M, 3F) | 21 (9M, 11F) | VH definition was based on consensus, with VH persistent for at least 3 months | 69.4; 66.2 | NR | MMSE 27.6; 28.2 | 62.2; 59.3 | 7.2; 7 (years) | 2.2; 1.8 | 22.5; 16.4 | LEDD 1031.2 mg; 805.2 mg | GDS 16.6; 15.5 | 7 stars |
| **↓Firbank et al. (2018),**<br>UK<br>Philips, 3.0 T | 17 (13M, 4F) | 19 (17M, 2F) | NPI (hallucination item) | 75.5; 72.3 | 11.6; 11.1 | MMSE 23.1; 25.6 | NR | 11; 9.6 (years) | NR | 55.9; 34.7 | Levodopa in 24h 717.3 mg; 673.5 mg | NR | 7 stars |
| **Functional Magnetic Resonance Imaging (fMRI)** |  |  |  |  |  |  |  |  |  |  |  |  |  |
| **↓Firbank et al. (2018),**<br>UK<br>Philips, 3.0 T<br>Visual task: checkboard paradigm | 17 (13M, 4F) | 19 (17M, 2F) | NPI (hallucination item) | 75.5; 72.3 | 11.6; 11.1 | MMSE 23.1; 25.6 | NR | 11; 9.6 (years) | NR | 55.9; 34.7 | Levodopa in 24h 717.3 mg; 673.5 mg | NR | 7 stars |
| **Knolle et al. (2020),**<br>Germany<br>Siemens, 3.0 T<br>Visual task: odd ball for salience processing | 14 (7M, 7F) | 23 (14M, 9F) | Presence of psychotic symptoms assessed with the CAARMS and the PANSS | 62.5; 63.1 | NR | MMSE 28; 29.4 | NR | 7.7; 9.9 (years) | 53.3% stage 1, 20% stage 2, 26.7% stage 3; 61.5% stage 1, 26.9% stage 2, 7.7% stage 3, 0% stage 4, 3.8% stage 5 | NR | Levodopa therapy (% yes) 80.8%; 86.7% | BDI 13; 8.1 | 7 stars |
| **Lefebvre et al. (2016),**<br>France<br>Philips, 3.0 T<br>Visual task: (Pins & ffytche, 2003) visual perception task | 18 (11M, 7F) | 16 (12M, 4F) | Presence of VH assessed with the NPI-C | 63.5; 62.69 | 12.44; 13.38 | MMSE 28; 28.88 | NR | 9.06; 8 (years) | 2; 2 | 25; 21.81 | LEDD 859.72 mg; 804.25 mg | Hamilton depression scale 3.56; 3.06 | 8 stars |
| **Stebbins et al. (2004),**<br>US<br>General Electric, 1.5 T<br>Visual task: stroboscopic vs. no visual stimulus, kinematic vs. stationary visual | 12 (NR) | 12 (NR) | Self-report on presence and frequency of VH. VH present at least three times per week. Severity was assessed with the NPI-Q and SAPS | 71.08; 73.25 | NR | MMSE 26.17; 27.96 | NR | 13.92; 11.17 (years) | 3; 3 | 30.42; 31.8 | NR | NR | 8 stars |
| **Meppelink et al. (2009),**<br>The Netherlands<br>Philips, 3.0 T<br>Visual task: detection task | 9 (5M, 4F) | 14 (11M, 3F) | Presence of complex VH at least weekly during the last month. Severity of VH assessed with the NPI | 61.2; 64.6 | 5.7; 5.7 | MMSE 26.8; 27.4 | NR | 8.1; 8.7 (years) | NR | 21.4; 20.4 | LEDD 855 mg; 794 mg | NR | 7 stars |

Neural substrates in PD psychosis
| Positron emission tomography (PET) |  |  |  |  |  |  |  |  |  |  |  |  |  |
| --- | --- | --- | --- | --- | --- | --- | --- | --- | --- | --- | --- | --- | --- |
| †Nishio et al. (2018), Japan NR | 19 (11M, 8F) | 53 (22M, 31F) | NPI | 69.4; 65.7 | 12.7; 11.2 | MMSE 26.6; 28.4 | NR | 7.3; 6.7 (years) | NR | 27.2; 16.2 | LEDD 612.4 mg; 436.4 mg | NR | 6 stars |
| †Nishio et al. (2018), Japan NR | 17 (7M, 10F) | 53 (22M, 31F) | NPI | 67.3; 65.7 | 13.4; 11.2 | MMSE 27.4; 28.4 | NR | 6.5; 6.7 (years) | NR | 18.7; 16.2 | LEDD 516.3; 612.4 mg | NR | 6 stars |
| †Nishio et al. (2018), Japan NR | 24 (12M, 12F) | 53 (22M, 31F) | NPI | 68.5; 65.7 | 13; 11.2 | MMSE 27.4; 28.4 | NR | 8.5; 6.7 (years) | NR | 23.1; 16.2 | LEDD 53 mg; 612.4 mg | NR | 6 stars |
| Park et al. (2013), South Korea Siemens, NR | 7 (3M, 4F) | 13 (8M, 5F) | VH assessed using the NPI administered by a neurologist | 71; 66.3 | 9.1; 9.2 | MMSE 26.1; 26.9 | NR | 5.4; 5.1 (years) | 2; 1.5 | 26.8; 22.1 | LEDD 537.1 mg; 531.4 mg | GDS 15.4; 13.3 | 7 stars |
| Boecker et al. (2007), Germany ECAT, EXACT HR +, CTI PET system | 8 (5M, 3F) | 11 (8M, 3F) | NPI (hallucination item) | 72.88; 70.56 | NR | MMSE 25.75; 26.82 | NR | 11; 8.05 (years) | 3.31; 2.68 | 46.25; 32.73 | Levodopa equivalent dose 667 mg; 617 mg | NR | 6 stars |
| Resting-state functional magnetic resonance imaging (rsfMRI) |  |  |  |  |  |  |  |  |  |  |  |  |  |
| ‡Yao et al. (2016), Hong Kong Philips, 3.0 T | 12 (10M, 2F) | 15 (10M, 5F) | Repetitive complex VH lasting for at least 4 weeks, and occurring at least once every 4 weeks | 69.38; 66.34 | 7.2; 6.3 | MMSE 27.625; 29 | NR | 9.1; 7.1 (years) | 3.1; 2.9 | 19.69; 20 | Levodopa 986.9 mg; 689.7 mg | MADRS-S 1.42; 0.63 | 7 stars |
| Shine et al. (2015), Australia General Electric, 3.0 T | 10 (NR) | 9 (NR) | Scores on the BPP | 69.5; 67.1 | NR | MoCA 26; 27.6 | NR | 6.9; 4.4 (years) | NR | 34; 32 | DDE (dopamine dose equivalent) 819.5 mg; 512.5 mg | BDI-II 15.5; 8.9 | 6 stars |
| Yao et al. (2014), Hong Kong Philips, 3.0 T | 12 (3M, 9F) | 12 (4M, 8F) | VH assessed with PPRS | 67.6; 63.4 | NR | MMSE 27.6; 28.5 | NR | 10; 8.4 (years) | 3.2; 2.8 | 20.9; 18 | LEDD 978.7 mg; 704.9 mg | NR | 7 stars |
| Bejr-Kasem et al. (2019), Spain Philips, 3.0 T | 18 (10M, 8F) | 14 (10M, 4F) | PD patients included if VH remained stable during the 3 months before inclusion in the study | 70.4; 65.8 | 12.5; 11.6 | PD-CRS, 91.9; 92.9 | NR | 5.2; 4 (years) | 2.1; 2.1 | 21.9; 25.8 | LEDD 697.2 mg; 601.1 mg | HADS-D 2.2; 3.3 | 7 stars |

Neural substrates in PD psychosis
| Single photon emission computed tomography (SPECT) |  |  |  |  |  |  |  |  |  |  |  |  |  |
| --- | --- | --- | --- | --- | --- | --- | --- | --- | --- | --- | --- | --- | --- |
| **Kiferle et al. (2014),**<br>Italy<br>General Electric,<br>NR | 18 (NR) | 18 (NR) | VH established with the NINDS-NIMH criteria | 75; 76.6 | NR | MMSE 24.9; 25.4 | 69.5; 70.8 | 66; 69.3 (months) | NR | 27.3; 25.8 | LEDD 547.3 mg; 496.2 mg | NR | 6 stars |
| **Jaakkola et al. (2017),**<br>Finland<br>General Electric,<br>NR | 22 (13M, 9F) | 48 (27M, 21F) | VH developed after baseline at follow up | 66; 64 | NR | NR | NR | 7; 6.7 (years) | 3.6; 3 | NR | LEDD 664 mg; 683 mg | NR | 8 stars |

†Studies that included one or more groups of PD psychosis patients with different symptoms, e.g., visual illusions, visual hallucinations.
‡Studies with more than one neuroimaging modality.
BDI: Beck Depression Inventory; BPP: Bistable Percept Paradigm; CAARMS: Comprehensive Assessment of At Risk Mental States; GDS: Geriatric Depression Scale; HADS-D: Hospital Anxiety and Depression Scale – depression item; HADS-A: Hospital Anxiety and Depression Scale – anxiety item; LEDD: Levodopa equivalent daily dose; MADRS: Montgomery-Asberg Depression Rating Scale; MDS-UPDRS: Movement Disorders Society Unified Parkinson's Disease Rating Scale; MMSE: Mini-Mental State Examination; MoCA: Montreal Cognitive Assessment; NPI: Neuropsychiatric Inventory; NPI-C: Neuropsychiatric Inventory – Clinician; NPI-Q: Neuropsychiatric Inventory – Questionnaire; NR: None reported; PANSS: Positive and Negative Symptom Scale; PD-CRS: Parkinson's Disease – Cognitive Rating Scale; PPRS: Parkinson Psychosis Rating Scale; VH: visual hallucinations.

All the 18 studies employed a cross-sectional design and employed different neuroimaging modalities: DTI (N=6), resting-state fMRI (N=4), PET (N=3), fMRI (N=5), and SPECT (N=2). All patients had idiopathic PD, the diagnostic tool or decision for a PD diagnosis involved mainly the UK Parkinson’s Disease Society Brain Bank criteria (Daniel & Lees, 1993; Hughes et al., 1992), whilst different criteria were used to assess the presence of psychosis symptoms: clinical rating scales (e.g., the Neuropsychiatric inventory (NPI); Movement Disorder Society Unified Parkinson’s Disease Rating Scale (MDS-UPDRS) part I item 1.2; Scales for Outcomes in Parkinson’s Disease– psychiatric complications (SCOPA-PC); Parkinson Psychosis Rating Scale; Bistable Percept Paradigm), and the National institute of Neurological Disorders and Stroke – National Institute of Mental Health (NINDS-NIMH) criteria (Ravina et al., 2007). The assessment of psychotic symptoms in PD patients mainly focused on visual hallucinations. The most common cognitive outcome measure used in these studies was the Mini-Mental State Examination (MMSE) (Folstein et al., 1975).

All studies had full score on the “Exposure” domain (i.e., ascertainment, non-response rate and ascertainment method for PDP and PDnP patients); “Comparability” was good in all studies due to their matched design, whereby patients were matched on age, gender and other clinical or demographic variables, whilst other studies reported these variables as covariates included in the analysis. Six studies received one star in this domain when only one factor, i.e., either age or other variable such as gender, was used as covariate or to match patients. “Selection” domain included selection and definition of both PDP patients (i.e., cases) and PDnP patients (i.e., controls), selection of PDP patients was assigned one star (i.e., the maximum) in all studies. Similarly, selection of PDnP patients was assigned one star in all studies, whilst definition of such group was clearly reported in five studies which were assigned one star in this domain. Full quality ratings are reported in eTable1 (Supplementary Material 2).

### 3.2. Descriptive analysis: Task-based functional MRI (fMRI) studies

Five case-control cross-sectional studies employed fMRI in conjunction with cognitive activation paradigms such as signal detection (Lefebvre et al., 2016; Meppelink et al., 2009), checkerboard paradigm (Firbank et al., 2018), oddball paradigm to examine salience (Knolle et al., 2020), and apparent motion (Stebbins et al., 2004). These studies included a total of 154 patients, of which 70 were PDP patients (52.9% male, mean age ± SD = 66.76 ± 6.22 years, mean education ± SD = 9.91 ± 3.67 years) and 84 were PDnP patients (62.8% male, mean age ±SD = 67.19 ± 5.16 years, mean education ± SD = 10.06 ± 3.94 years). One study applied a visual pop-out task whereby patients were asked to press a button when they recognised an image appearing from a background noise (Meppelink et al., 2009). They examined brain activation before and at the moment of the pop-out condition of the task using an ROI approach. Before the pop-out, activation of bilateral parietal cortex and occipital and frontal cortices with a lower threshold were reported PDP patient with VH (n=9), whilst middle occipital cortex and inferior frontal gyrus were reported in PDnP patients (n=14). When these activation patterns were compared between groups based on the time before detection of the stimulus, PDP patients with VH showed decreased activation in bilateral occipital cortex (at -5.8 to -4.6 s), superior frontal gyrus (at -3.5 to – 2.3 s), bilateral fusiform gyrus and left lingual gyrus, cingulate cortex and right middle frontal gyrus (at -1.2 s) compared to PDnP patients. At the moment of the pop-out, there was no significant difference in brain patterns between PDP and PDnP patients. Similarly, a signal detection task was also used to test visual processing, patients were asked to say whether or not they saw a grey-scale grid appearing on a background (Lefebvre et al., 2016). PDP patients with minor VH (n=18) showed hyperactivation at detection of the stimulus in the right cerebellum, right prefrontal cortex and right occipital cortex, whilst hypoactivation of the left cingulate cortex, caudate nucleus, temporal and occipital cortices compared to PDnP patients (n=16); the analyses were corrected at cluster level. Similarly, aberrant brain activation was also found by Stebbins et al. (Stebbins et al., 2004) who applied moving and stroboscopic images to assess basic and apparent motion visual perception using whole-brain approach. PDP patients (n=12) reported greater activation in superior frontal lobe, and caudate nucleus compared to non-hallucinating PDnP patients (n=12). Conversely, this latter group reported greater activation of temporal-occipital lobe, supramarginal parietal lobe, inferior parietal lobe, and cingulate cortex compared to the former. Decreased the activation of V5/MT area in the former group was also observed during such condition compared to the latter; however, this was detected with a small volume correction analysis. When stroboscopic images were presented, PDP patients had increased activation in inferior frontal lobe and caudate nucleus compared to PDnP patients. The latter group showed greater activation in other areas, namely cingulate cortex and inferior parietal lobe. In contrast, Firbank et al. (Firbank et al., 2018) did not find any differences in V5 activation and any other brain region between their patient groups. They applied a visual task based on a checkboard paradigm (previously described by Taylor et al. (Taylor et al., 2012)) during which patients were asked to passively observe the flickering checkerboard and to press a button when a pink dot appeared on screen. Only one study examined salience network in PDP. Knolle et al. (Knolle et al., 2020) examined salience processing by using an oddball visual task whereby patients were presented with alternating images of faces and outdoor scenes, these were either neutral stimuli or emotionally charged (e.g., angry face or image of a car crash). They measured cerebral blood flow and blood-oxygen level dependent (BOLD) signals in four ROIs: substantia nigra (SN)/ventral tegmental area (VTA), ventral and dorsal striatum, bilateral hippocampus, and bilateral amygdala. They observed hyperactivation to emotionally salient stimuli in PDP patients (n=14) in the amygdala, hippocampus, striatum and SN/VTA compared to PDnP patients (n=23). They also assessed novelty salience in addition to emotional salience by presenting new neutral stimuli in their paradigm. However, there were no differences in brain activation between groups in the novel stimuli condition (Knolle et al., 2020).

Collectively, task-related decrease in activation in the occipital-parietal-frontal (Meppelink et al., 2009) and temporal regions and subcortical structures such as the cingulate cortex and caudate (Lefebvre et al., 2016) as well as increase in activation of the frontal cortex (Lefebvre et al., 2016; Stebbins et al., 2004) and subcortical areas (Knolle et al., 2020) was observed in PDP patients compared to PDnP patients (Fig. 2).

**Fig. 2.**
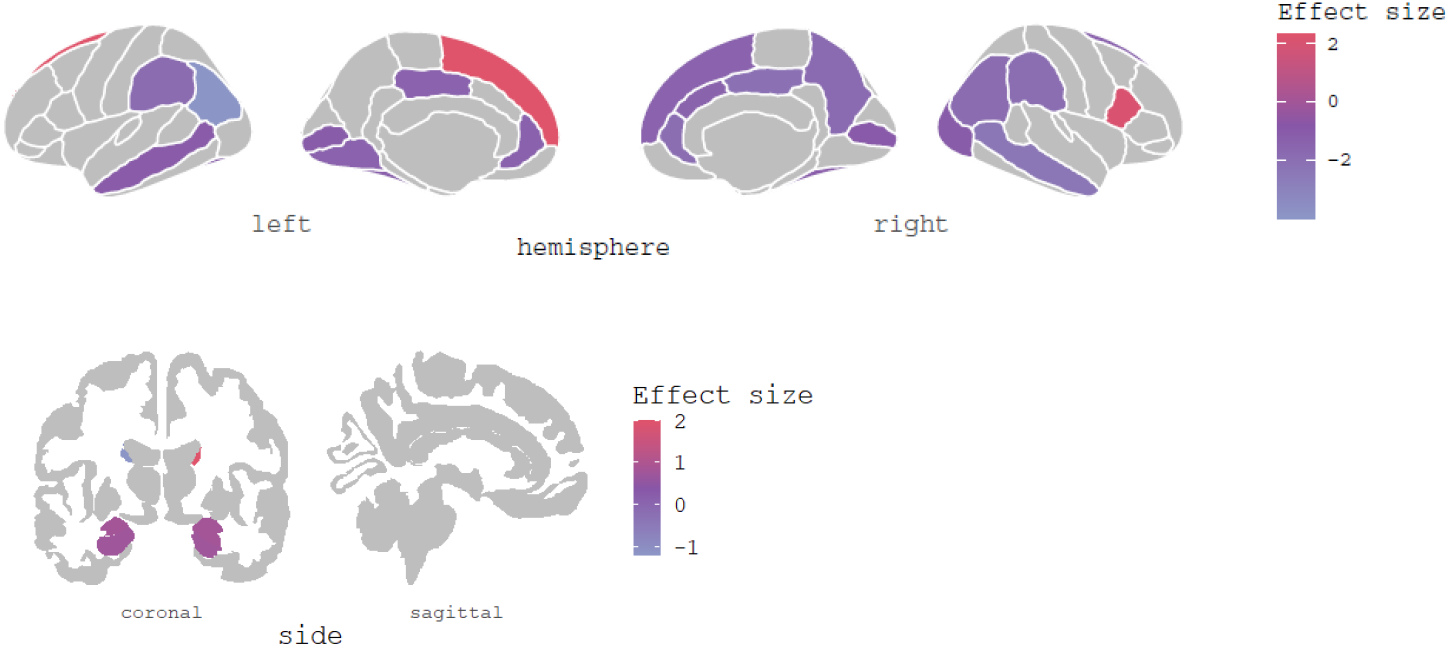
Plots displays the brain regions showing hyper-(red) and hypo-(violet) activations across all fMRI studies (N=5), where data were available. Z scores and/or t values were extracted from the studies and converted into effect size estimates for purely display purposes (without carrying out any quantitative synthesis).

### 3.3. Descriptive analysis: Resting state fMRI (rsfMRI) studies

Two studies examined the Default Mode Network (DMN) and its nodes (Bejr-Kasem et al., 2019; Yao et al., 2014), one investigated the hippocampus (Yao et al., 2016), and one study focused on attentional networks (Shine et al., 2015) using resting-state fMRI. These studies included a total of 102 patients, of which 52 were PDP patients (44.2% male; mean age ± SD = 69.38 ± 1.28 years, mean education ± SD = 9.85 ± 3.75 years) and 50 were PDnP patients (48% male; mean age ± SD = 65.58 ± 1.56 years; mean education ± SD = 8.95 ± 3.75 years). In a sample of 32 PDP patients (18 with minor VH), Bejr-Kasem et al. (Bejr-Kasem et al., 2019) reported differences in functional connectivity in the posterior cingulate cortex (PCC), one of the nodes in the Default Mode Network (DMN) between patient groups. Patients did not differ in age, gender, years of educations, and cognitive impairments. The PCC was used as seed of interest to investigate the co-activation at rest between the DMN and other attentional networks. In PDP patients with minor VH, the PCC showed greater connectivity with bilateral middle temporal gyrus (i.e., DMN area) and bilateral superior parietal lobes, right precentral gyrus, and left middle cingulate cortex, regions involved in task-based attention-demanding activities. Results showed greater connectivity in PDP patients with minor VH between PCC and left middle occipital gyrus and bilateral posterior middle temporal gyrus, which are involved in visual information processing. When alternative seeds were applied and grey matter volume of PCC was entered as covariate, results did not change, suggesting that these patterns of connectivity are inherent and independent of PCC atrophy. A similar pattern of findings was also reported by Yao et al. (Yao et al., 2014) who examined the DMN in 24 PDP patients of which 12 reported VH using probabilistic independent component analysis (ICA) and dual regression. Patients between the two groups did not differ in age, gender, and cognitive impairments (assessed with MMSE). Across 40 components identified by the probabilistic ICA temporally correlated within the DMN, in PDP patients with VH greater functional co-activation in the right middle frontal gyrus and bilateral posterior cingulate gyrus and precuneus was reported compared to PDnP patients. This co-activation was not associated with severity of psychosis symptoms. One study examined the temporal activation of different attentional networks in the two patient groups. Shine et al. (Shine et al., 2015) applied a bistable percept paradigm to define group allocation to PD patients with VH (n=10) or without VH (n=9). Patients also underwent a visual task which assessed the strength of mental imagery in both groups. This was defined as the ability to recall a clear and vivid stimulus without the stimulus being physically present (Shine et al., 2015). The authors investigated the relationship between the strength of such ability and the co-activations of different attentional networks at rest. Clusters within the dorsal attention network (bilateral superior parietal lobule, bilateral frontal eye field), the DMN (midline precuneus, midline medial prefrontal cortex, bilateral hippocampal formation), the ventral attention network (bilateral anterior insula, bilateral dorsal anterior cingulate cortex), and the visual network (bilateral occipital cortex) were selected as ROI. The results showed that greater degree of mental imagery was positively associated with activation of the ventral attentional network and the DMN. Both networks are associated with internal attention processing. High mental imagery was associated with decreased connectivity between dorsal and ventral attentional networks, and dorsal attentional and visual networks. The authors also examined the performance on the bistable percept paradigm and its relationship with the connectivity within these networks. Results showed that worse performance on this task was predictive of increased connectivity in the ventral attentional network and the DMN, and between the ventral and dorsal attention network. Mental imagery was not predictive of such relationship in these networks; however, it was associated with dysfunctions in connectivity between ventral attentional and visual networks. Compared to these three studies, Yao et al. (Yao et al., 2016) used the hippocampus as seed to examine its temporal co-activation with the whole brain. Differential patterns of hippocampal co-activation were reported for PDP patients with VH (n=12) and PDnP patients (n=15). The latter showed reduced hippocampal functional connectivity with parietal and frontal regions. Conversely, PDP patients with VH reported reduced functional co-activation between right hippocampus and bilateral cuneus and lingual gyrus, right fusiform gyrus, right medial temporal lobe and left superior/middle temporal gyrus; reduced functional connectivity between left hippocampus and bilateral lingual gyrus, right fusiform gyrus, left cuneus, right medial temporal lobe and right precuneus. PDP patients with VH also showed greater hippocampal functional connectivity with bilateral frontal lobe, cingulate cortex, and inferior parietal lobe compared to PDnP patients. In all analyses, age and cognitive impairments as measured with the MMSE were used as covariates. Visual accuracy was also controlled in the analysis. PD severity, LEDD, and gender were not correlated with functional connectivity. However, functional connectivity within the right occipital gyrus and right medial temporal lobe was correlated with cognitive scores of a visuospatial memory test, whereby lower scores on the test were associated with higher mean functional connectivity in both clusters.

Overall, greater functional connectivity between the Default Mode Network (DMN) nodes (Yao et al., 2014) and between the DMN and occipital-parietal-temporal areas (Bejr-Kasem et al., 2019) were observed in PDP compared to PDnP patients. Increased connectivity within dorsal attentional and ventral attentional networks, DMN, and visual network was also reported in PDP compared to PDnP patients (Shine et al., 2015). Opposite patterns of functional connectivity between bilateral hippocampus and frontal, parietal and temporal areas were also reported in PDP compared to PDnP patients in another study (Yao et al., 2016) (Fig. 3A-D).

**Fig. 3.**
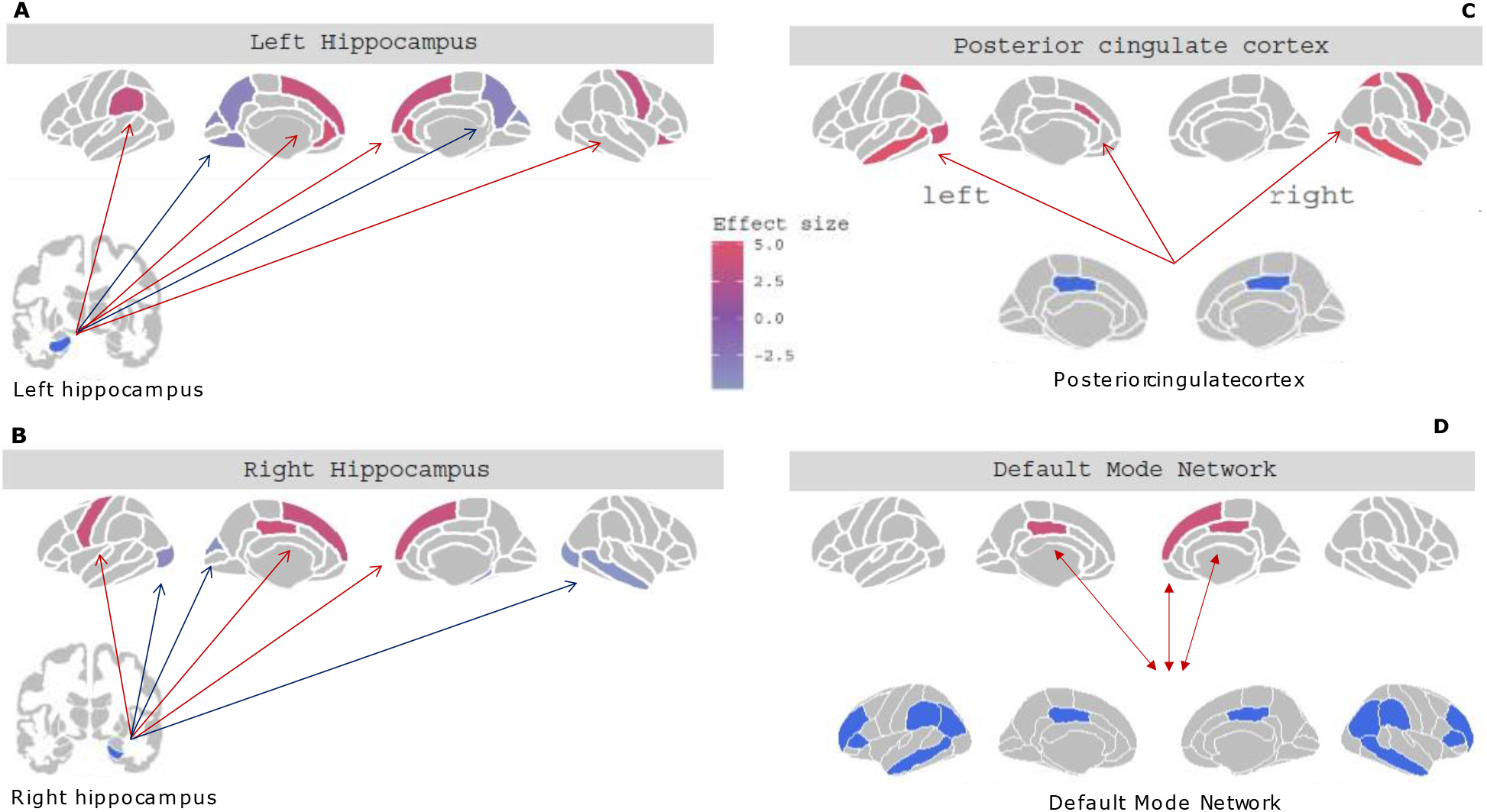
Patterns of functional connectivity of different brain areas. Specifically, regions functionally connected with the left (A) and right (B) hippocampi, with the PCC (C), and within the DMN (D). Red arrows connecting the regions represent greater connectivity in PD psychosis patients compared to PD patients, blue arrows represent decreased connectivity in PD psychosis patients compared to PD patients. Brain regions highlighted in dark pink-magenta show increased functional co-activation, brain regions highlighted in light purple show decreased functional co-activation. T values were extracted and used as effect size estimates from the studies and used purely for display purposes. Figures were created using *ggseg* (Mowinckel & Vidal-Piñeiro, 2020) in R (Team, 2013). These plots do not reflect any quantitative analyses conducted on these studies.

### 3.4. Descriptive analysis: Diffusion tensor imaging (DTI) studies

Six case-control studies used DTI to assess white matter tracts in PD psychosis. A total of 273 patients of which 83 were PDP with visual hallucinations (65.1% males: mean age ± SD = 69.42 ± 3.49 years; mean education ± SD = 11.967 ± 4.96 years) and 190 were PDnP patients (57.4% males; mean age ± SD = 67.12 ± 2.69 years; mean education ± SD = 11.43 ± 5.31 years) were analysed. Three studies applied an ROI approach (Hepp et al., 2017; Lee et al., 2016; Yao et al., 2016), two applied tract-based spatial statistics (TBSS) (Firbank et al., 2018; Lee et al., 2017), and one used different analytical approaches such as anatomically constrained tractography and network-based statistics (Zarkali et al., 2020). Hepp et al. (Hepp et al., 2017) examined the nucleus basalis of Meynert (NBM), a cholinergic sub-structure located in the basal forebrain. They examined whole-brain fibre tracts between PDP patients with VH (n=15) and PDnP patients (n=40), both comparable in demographics and clinical variables, and both de novo patients with idiopathic PD. No differences in mean diffusivity (MD) or fractional anisotropy (FA) between groups in tracts from the NBM and frontal, temporal, cingular, and insular regions were observed. However, group differences were found in parietal and occipital tracts, whereby PDP patients showed higher MD in these two tracts compared to PDnP patients thus suggestive of microstructural damages in parietal and occipital tracts. Lee et al.(Lee et al., 2016) investigated four main regions: the lateral geniculate, the optic chiasm, the optic radiation, and the optic nerve. Patients did not differ in age, gender, and PD symptoms. PDP patients (who experienced VH, n = 10) showed alterations of all the regions investigated compared to PDnP patients (n=14), however MD and AD were significantly higher in the left optic radiation and right optic nerve in PDP patients respectively. The lateral geniculate nucleus showed mildly reduced volume in this patient group compared to PDnP patients. These findings are in line with the above study suggesting fibre tracts and white matter abnormalities in visual processing areas in PDP. Only one study examined the hippocampus focusing on MD and voxel-by-voxel approach. Yao et al. (Yao et al., 2016) observed higher MD in the right hippocampus and right posterior hippocampal regions in PDP patients with VH (n=12) compared to PDnP patients (n=15). This was not associated with LEDD, gender and PD severity. However, TBSS studies revealed opposite pattern of results. Firbank et al. (Firbank et al., 2018) showed no significant differences in MD in PDP patients (n=17) compared to PDnP patients (n=18). Lee et al. (Lee et al., 2017) reported widespread white matter abnormalities across the cortex but these were not significantly different between PDP patients (n=10) and PDnP patients (n=21). Zarkali et al. (Zarkali et al., 2020) revealed structural connectivity dysfunctions which were widespread in PDP patients with VH (n=19) compared to PDnP patients (n=81). They were matched in age, gender, motor scores and cognitive impairments. They applied a different approach to study white matter changes, network-based statistics and analysis of controllability which enables to estimate the impact of cortical changes on brain functions. Their results showed a network of 82 nodes in PDP patients with VH that corresponds to regions involved in information processing and attention; their analysis showed reduced structural connectivity in this network. They also examined regional gene expression in areas that were affected by these structural changes and, using enrichment analysis, found dysfunctions in mRNA and chromosome metabolism in these areas. This suggests abnormalities at a cortical and more intrinsic levels involve molecular processes in PDP patients with VH.

Abnormalities in parietal, occipital (Hepp et al., 2017) and hippocampal tracts (Yao et al., 2016), optic radiation and optic nerve (Lee et al., 2016), and across the nodes involved in information processing and attention (Zarkali et al., 2020) were reported in PDP compared to PDnP patients. However, two studies did not report differences in white matter tracts between groups (Firbank et al., 2018; Lee et al., 2017) (Fig. 4).

**Fig. 4.**
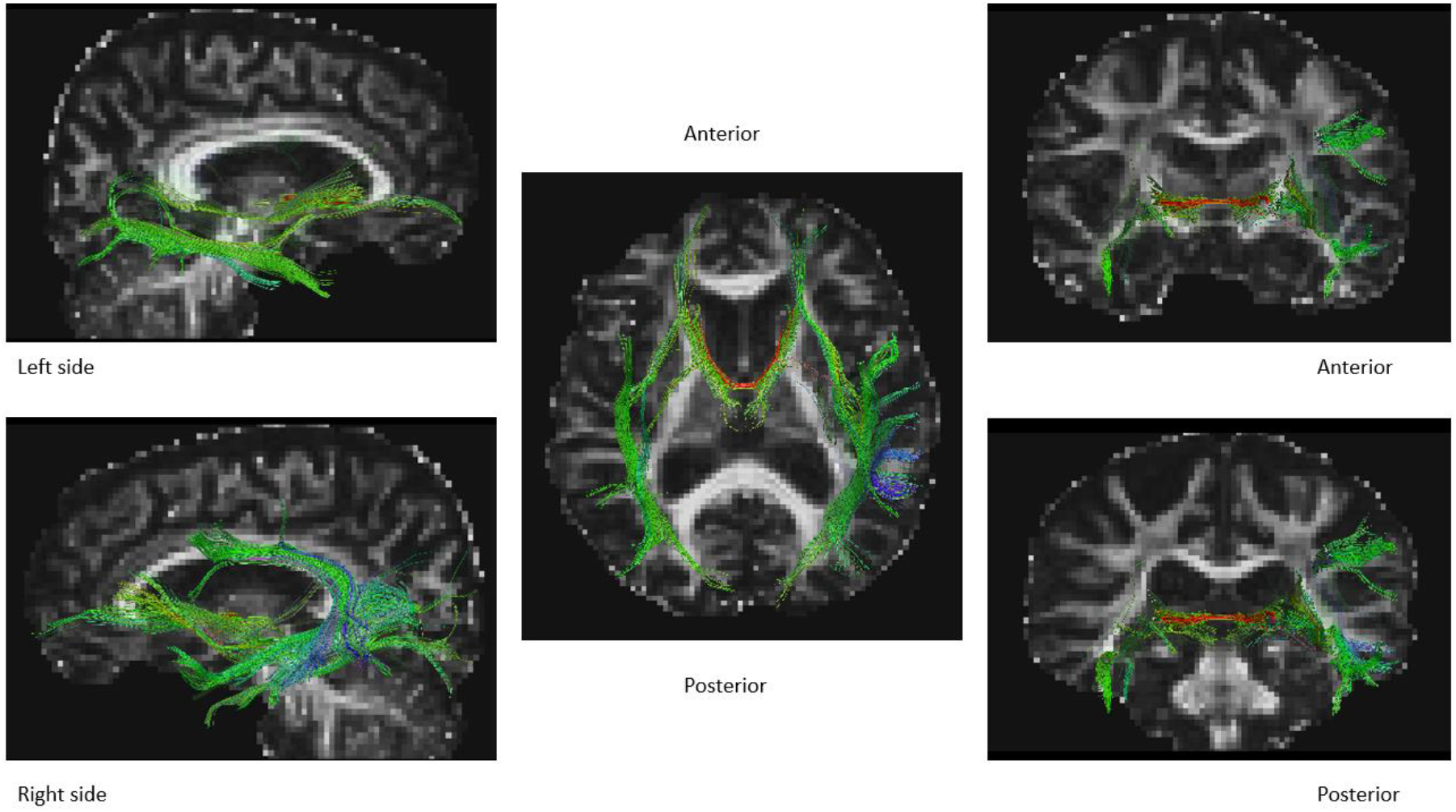
White matter fibre tracts that were found dysfunctional in PDP patients compared to PDnP patients in DTI studies (n=4). Information was extracted from each paper and plotted on a brain template on Trackviz (Wang et al., 2007). These plots do not reflect any analyses conducted on these studies and is purely for display purposes.

### 3.5. Descriptive analysis: Positron emission tomography (PET) studies

Three case-control studies employed PET imaging to examine glucose metabolism in PDP patients. A total of 152 patients of which 75 were PDP with visual hallucinations (50.6% males; mean age ± SD = 69.52 ± 2.40 years; mean education ± SD = 12.05 ± 1.99 years) and 77 were PDnP patients (49.4% males; mean age ± SD= 67.52 ± 2.65 years; mean education ± SD = 10.20 ± 1.41 years) were analysed, and all studies employed an 18-F -FDG PET. One study (Nishio et al., 2018) divided PD patients according to their visual symptoms, indicative of PDP: 19 had visual hallucinations, 17 experienced visual illusions and 24 reported kinetopsia which involves a motion misperception of a stationary object. PDP patients with VH were significantly older, had more cognitive decline, had more severe motor symptoms, and were administered higher LEDD than PDnP patients. In this study, the authors applied a partial least square correlation analysis and examined glucose metabolism in PDP patients with different visual symptoms. Presence of VH and kinetopsia in PD patients was associated with hypometabolism in temporo-parietal regions, whilst presence of visual illusions was associated with hypometabolism in occipital and temporo-parietal metabolism. Thus, suggesting deficits in visual processing areas in PDP, specifically if considering visual illusions as psychotic symptom. One early study also supports this finding. Boecker et al. (Boecker et al., 2007) examined PDP patients with VH (n=8) and PDnP patients (n=11), patient group did not differ in age, gender and LEDD. They found differences in regional cerebral metabolic glucose consumption rate between groups in left occipital, temporal and parietal areas in PDP patients with VH (corrected for false discovery rate). Specifically, left inferior parietal lobule, left supramarginal gyrus, bilateral precuneus, right inferior parietal lobule, right cingulate cortex, left middle frontal gyrus and left middle temporal gyrus. These areas are considered part of the dorsal visual pathway. Left parahippocampal gyrus and left lingual gyrus were also found to have low glucose consumption rate in this group. These two areas are associated with ventral visual pathway, thus complementing previous findings of deficits in visual processing in people with PDP (Collerton et al., 2012; Weil et al., 2016). A third study also examined cognitive impairments in PDP patients with VH (n=15) compared to PDnP patients (n=13) (Park et al., 2013). In a first group comparison, PD patients with VH showed hypometabolism bilaterally in middle and inferior temporal cortices, left lingual gyrus, left angular gyrus compared to PDnP patients. Within PD patients with VH, those with cognitive impairment showed reduced metabolism in middle and inferior temporal cortices, bilateral frontal areas, and left superior parietal and occipital regions; this was uncorrected with a threshold of *p* < 0.01. Reduced metabolism in the precuneus, bilaterally and superior parietal areas as well as in frontal, temporal and occipital lobes was observed (uncorrected, *p* < 0.001) in PD patients with VH and cognitive deficits compared to those without cognitive impairments. Correlation analysis showed that lower scores on the MoCA (Nasreddine et al., 2005) were associated with hypometabolism in bilateral superior parietal and middle occipital gyri, left precuneus, and right fusiform gyrus. Although these findings support previous results, this study is also suggestive of a more widespread pattern of altered glucose metabolism involving temporal, occipital, and parietal cortices. Overall, general patterns of hypometabolism were observed in occipito-temporo-parietal regions (Nishio et al., 2018) including regions within the dorsal and ventral visual pathways (Boecker et al., 2007). PDP patients also showed reduced metabolism in bilateral frontal, temporal and occipital cortices (although uncorrected) compared to PDnP patients irrespective of presence or absence of mild cognitive impairment (Park et al., 2013) (Fig. 5).

**Fig. 5.**
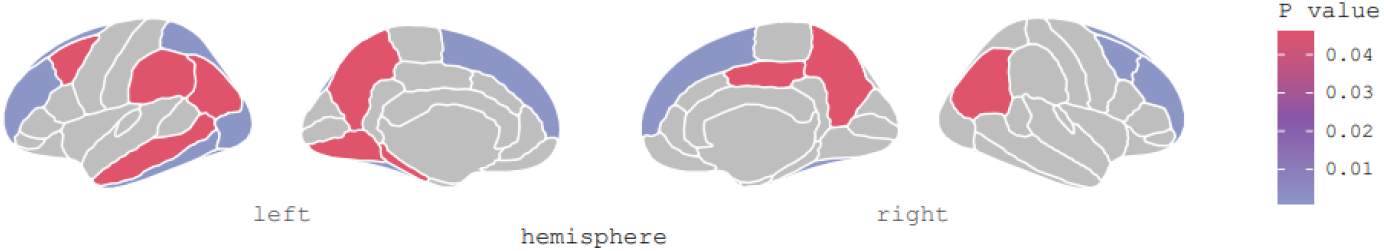
Plot shows brain regions with hypometabolism in PDP patients compared to PDnP patients. P values from the studies were extracted and used for display purposes without carrying out any quantitative synthesis. All figures were created using *ggseg* (Mowinckel & Vidal-Piñeiro, 2020) in R.

### 3.6. Descriptive analysis: Single-photon emission computerised tomography (SPECT) studies

Only two studies employed a [123 I] FP-CIT SPECT scan which is applied to examine dopamine transporter (DAT) availability in specific brain regions. Jaakkola et al. (Jaakkola et al., 2017) examined longitudinally a cohort of PD patients who then developed psychosis symptoms. Although there was no difference in socio-demographics and in PD medications between patients who transition to PDP, data revealed lower DAT binding at baseline in right and left ventral striatum and right putamen. They also measured binding ratio in specific ROIs: bilateral caudate, bilateral anterior putamen, and bilateral posterior putamen, using occipital region as reference. They observed that decreasing binding ratio in ventral striatum bilaterally and right putamen in PDP patients compared to PDnP patients. Asymmetry in putamen in PDP patients was also observed. At follow-up, PDP patients had reduced DAT binding in the right amygdala compared to PDnP patients. Similarly, Kiferle et al. (Kiferle et al., 2014) retrospectively analysed data from PD patients (n=36) who then developed psychosis (n=18) at 72.8 months from PD onset (61.5 months after starting PD medications). Reductions in DAT binding in right caudate in PDP patients were reported at baseline compared to PD patients who did not develop such symptoms; no significant differences in cognitive state, PD symptoms and LEDD were reported between these two groups. Overall, PDP patients reported reduced DAT binding in right caudate at baseline (Kiferle et al., 2014), lower DAT binding in right and left ventral striatum, and right putamen, in addition to decreased binding ratio in ventral striatum bilaterally and right putamen (Jaakkola et al., 2017) compared to PDnP patients (Fig.6).

**Fig. 6.**
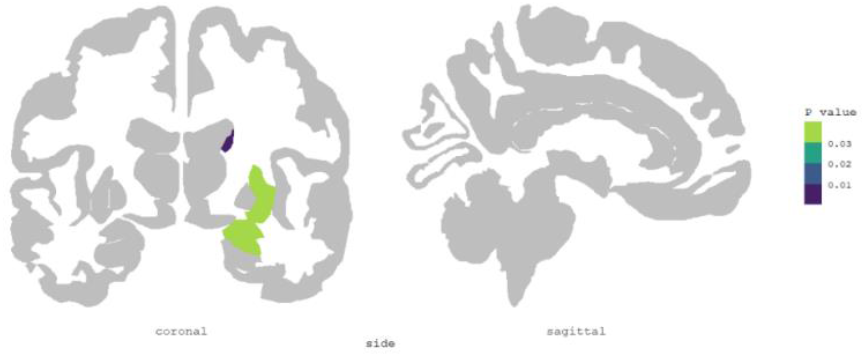
Pattern of abnormal dopamine transporter (DAT) binding in PD psychosis patients P values were extracted and used as effect size estimates from the studies, for purely display purposes.

## 4. Discussion

As stated earlier, here we synthesised evidence from neuroimaging studies across multiple modalities using a qualitative approach with a view to identify key brain alterations that may be implicated in PDP. Findings from task-based fMRI studies revealed decreased activity across the cerebal cortex in PDP compared to PDnP patients, mainly localised in temporal, parietal and occipital lobe. Some areas of hyperactivation were observed in the frontal lobe but with few studies reporting such increased activation (Knolle et al., 2020; Lefebvre et al., 2016). Alterations in brain activity found in task-based fMRI studies varied according to the cognitive paradigm employed in the studies. While no consistent pattern of functional alteration was observed across the different cognitive paradigms this may reflect both differences in cognitive processes engaged by these paradigms as well as analytic approaches that included whole-brain or ROI analysis approaches. Evidence from resting-state fMRI studies more commonly reported patterns of increased connectivity of frontal, parietal and temporal areas in PDP patients. Although employing different approaches (e.g., seed-to-voxel and region of interest), these results showed increased connectivity at rest across areas involved in attentional networks. Using different seeds, e.g., posterior cingulate cortex, bilateral hippocampus, and the DMN, frontal regions appear to show more functional connectivity with these seeds in PDP compared to PDnP patients. Results from DTI studies present evidence which suggests additional areas with abnormalities in PDP patients, that is microstructural damages and white matter abnormalities, expressed as higher mean diffusivity, in occipital, parietal tracts, and areas involved in visual processing (Hepp et al., 2017; Lee et al., 2016) as well as dysfunctions in hippocampal tracts (Yao et al., 2016). However, two DTI studies reported no difference between patient groups (Firbank et al., 2018; Lee et al., 2017). Consistent patterns of hypometabolism were reported in PDP across all studies (n=3) in occipito-temporo-parietal areas as well as frontal cortices when cognitive impairments were controlled for in the analysis. Reduced DAT binding in sub-cortical regions in PDP was reported in both SPECT studies. Collectively, these findings suggest a widespread network of functional, metabolic, and white matter dysfunctions in PDP. These were evident primarily in occipito-parietal-temporal regions in PDP compared to PDnP patients, thus in agreement with previous evidence (Ffytche et al., 2017; Lenka et al., 2020; Lenka et al., 2015; Pisani et al., 2022). These areas are part of the dorsal and ventral visual pathways, involved in higher order visual processing which may suggest a more prominent role of visual information processing in the manifestation of PD psychosis. We also observed dysfunctions in nodes within the DMN, and attentional networks which further aligns with previous evidence suggesting a possible overreliance on self-referential and internal endogenous processes (i.e., “top-down”), due to disengagement or abnormal functioning of dorsal and ventral attentional network giving rise to hallucinations in PD patients (Hall et al., 2016; Hall et al., 2015; Muller et al., 2014; O’Callaghan et al., 2017; Shine et al., 2012; Shine, Halliday, et al., 2014; Shine, O’Callaghan, et al., 2014). Although occipital, temporal and parietal areas appear to be most affected in PD psychosis patients, evidence from PET and functional studies (both task-based and resting-state fMRI) also revealed patterns of dysfunction in frontal and sub-cortical cortices (i.e., frontal gyri, cingulate cortex). These areas are involved in executive and cognitive functions, such as selective attention, cognitive flexibility, inhibition (frontal cortices) (Alvarez & Emory, 2006; Stuss, 2011), and internally directed thoughts, arousal, cognitive control and emotional processing (cingulate cortex) (Allman et al., 2001; Bush et al., 2000; Leech & Sharp, 2014). Abnormalities within these areas may further contribute to the rise of psychotic symptoms in PD and may indicate that a wider cortical network involved in sophisticated executive functions may be also responsible for these symptoms. Furthermore, evidence from DTI and resting-state fMRI studies here reviewed appear to also suggest a possible role of the hippocampus (both as seed of interest and as part of the DMN) in PDP showing microstructural damages in posterior hippocampus, and abnormal patterns of functional co-activation in bilateral hippocampi (Shine et al., 2015; Yao et al., 2016) with parietal, temporal and frontal areas. This is also aligned with the findings from the mega-analysis by Vignando et al. (Vignando et al., 2022) which reported grey matter volume reduction in hippocampus bilaterally, after covarying for age, gender, disease onset, PD severity, medication, and cognition, suggesting a role of this area independent of cognitive decline in PD psychosis. This may suggest that development of psychosis in PD patients may be due to cortical volume loss in certain nodes, e.g., hippocampus, consequently leading to abnormal functionality at a network level. However, more research is needed to understand the potential temporal relationship between cortical volume loss and abnormal activation across the cortex. Future studies should also be carefully tailored to investigate the role of the hippocampus in this clinical population.

### 4.1 Limitations and strengths

We were unable to quantitatively synthesise data from different neuroimaging modalities due to the numerous statistical methodologies employed, e.g., ROI vs. whole-brain, seed-to-voxel vs. network analysis, and the limited number of studies using PET and SPECT. Therefore, drawing definitive conclusions from these findings may be challenging. Similarly for DTI studies, we were unable to meaningfully analyse their results due to the different techniques employed, e.g., TBSS (Firbank et al., 2018; Lee et al., 2017) vs. ROI (Hepp et al., 2017; Lee et al., 2016; Yao et al., 2016) vs. network-based statistics (Zarkali et al., 2020). We were also unable to relate patterns of abnormal activity or connectivity with different types of psychotic symptoms as an insufficient number of studies reported these. Future studies need to examine the association between the wide spectrum of psychosis manifestations in PD, such as presence/passage, auditory and multimodal hallucinations as well as delusions, to shed further light on common pathways shared by different types of symptoms. In addition, we noticed a prominent use of visual tasks in task-based fMRI studies specifically targeting general visual information processing. One study (Knolle et al., 2020) used a visual task in the context of emotional salience which suggested a role of sub-cortical areas in processing emotionally charged images in PD psychosis. Further studies should consider employing a variety of fMRI tasks to examine for example the activation of memory-related areas (e.g., hippocampus), and other cognitive domains (e.g., working memory, executive functions). These can further expand current knowledge on the neural correlates contributing to the development of psychosis in PD patients. We did not find studies using MRS approach in this clinical population. This is quite interesting, MRS modality can provide helpful information on local concentration of specific chemicals (e.g., glutamate and GABA) in specific cortical and sub-cortical areas. Such modality can shed further lights on the neurobiological mechanisms possibly involved in PD psychosis. Lastly, this is a systematic synthesis of different studies, and therefore aimed to provide a qualitative framework to better understand the neural substrates implicated in PD psychosis. We attempted to synthesise the results from different studies using graphical representations of the brain areas involved in PD psychosis by employing *ggseg* package (Mowinckel & Vidal-Piñeiro, 2020). This enabled us to visually represent patterns of abnormal activity and metabolism, and connectivity in PDP patients compared to PDnP patients. We also employed Trackviz (Wang et al., 2007) to visualise fibre tracts that were dysfunctional in PDP patients. This can provide initial evidence to conduct future meta-analysis using quantitative approach to examine each neuroimaging modality.

### 4.2. Conclusion

In conclusion, the qualitative synthesis revealed extensive dysfunction in PD psychosis patients, ranging from hypoactivation in task-based fMRI and abnormal functional connectivity in resting-state fMRI studies, hypometabolism and microstructural damages in this clinical population. There is initial evidence also suggesting a role of parietal, temporal and occipital regions involved in higher order visual processing, and endogenous/exogenous attentional networks, as well as sub-cortical areas, such as the hippocampus. These findings may reflect the preponderance of PD patients with visual hallucinations investigated in these studies.

## Data Availability

Data availability statement: This publication is a systematic review, and as such it includes data that have already been published. The data that support the findings of this study are available from the corresponding author upon request.

## Acknowledgment

None

## Authors’ contributions

*Conceptualisation:* Sara Pisani, Latha Velayudhan, Sagnik Bhattacharyya

*Methodology:* Sara Pisani, Brandon Gunasekera, Latha Velayudhan, Sagnik Bhattacharyya

*Investigation:* Sara Pisani, Latha Velayudhan, Sagnik Bhattacharyya

*Data curation:* Sara Pisani, Yining Lu

*Formal analysis:* Sara Pisani, Brandon Gunasekera, Yining Lu, Sagnik Bhattacharyya

*Visualisation*: Sarra Pisani, Sagnik Bhattacharyya

*Funding acquisition*: Sagnik Bhattacharyya, Latha Velayudhan, Dominic ffytche, Dag

Aarsland, Kallol Ray Chaudhuri, Clive Ballard

*Writing (original draft):* Sara Pisani, Sagnik Bhattacharyya, Latha Velayudhan

*Writing (review & editing):* All authors

*Supervision*: Latha Velayudhan, Dominic ffytche, Sagnik Bhattacharyya

